# Can a specific biobehavioral based therapeutic education program lead to changes in pain perception and brain plasticity biomarkers in chronic pain patients? A study protocol for a randomized clinical trial

**DOI:** 10.1101/2023.07.19.23292903

**Authors:** Silvia Di Bonaventura, Josué Fernández Carnero, Raúl Ferrer Peña

**Affiliations:** Escuela Internacional de Doctorado, Department of Physical Therapy, Occupational Therapy, Rehabilitation and Physical Medicine, Rey Juan Carlos University, 28933 Alcorcón, Spain; Department of Physical Therapy, Occupational Therapy, Rehabilitation and Physical Medicine, Rey Juan Carlos University, 28922 Alcorcón, Spain; Cognitive Neuroscience, Pain and Rehabilitation Research Group (NECODOR), Faculty of Health Sciences, Rey Juan Carlos University, Madrid, Spain; Grupo Multidisciplinar de Investigación y Tratamiento del Dolor, Grupo de Excelencia Investigadora URJC-Banco de Santander, 28922 Madrid, Spain; La Paz Hospital Institute for Health Research, IdiPAZ, 28029 Madrid, Spain; Motion in Brains Research Group. Centro Superior de Estudios Universitarios La Salle, Universidad Autonóma de Madrid, 28023 Madrid, Spain; Grupo de Investigación de Dolor Musculoesqueletico y Control Motor, Universidad Europea de Madrid, 28670 Villaviciosa de Odón, Spain; Departamento de Fisioterapia, Facultad de Ciencias de la Salud, CSEU La Salle, Universidad Autonóma de Madrid, 28023 Madrid, Spain

**Author notes:** Sponsor contact information:* Raúl Ferrer Peña. Sponsor and funder:* CSEU La Salle (UAM), Madrid. Committees:* Rey Juan Carlos University Ethical Committee.

**Keywords:** pain neuroscience education, brain derived neurotrophic factor, persistent pain, physical therapy.

## Abstract

**Abstract:** *Background:* Chronic pain conditions are complex multifactorial disorders with physical, psychological, and environmental factors contributing to their onset and persistence. Among these conditions, the role of brain-derived neurotrophic factor (BDNF) and the impact of a specific therapeutic education (TE) on pain management have emerged as important areas of research.

*Objective:* The aim of this study is to investigate the effects of a specific type of therapeutic education on pain levels and BDNF concentrations.

*Methods:* In this single-blind, randomized clinical trial, patients will be randomly assigned to one of two groups: one group will receive education with TE and the other group will receive education without TE. Assessments will be made at baseline, mid-treatment, post-intervention, and at one and eight months.

*Outcomes:* The expected results of this study will shed light on the role of TE in pain management and provide insight into its effects on BDNF levels. The intervention will also provide an opportunity to biochemically measure the plastic changes, thereby improving our understanding of its effects on brain plasticity.

*Conclusion:* This study aims to determine the applicability of therapeutic education as an effective non-pharmacological intervention for the management of chronic pain. The rigorous scientific methods used ensure that the proposed interventions will be clinically applicable across different health care systems.

*Strengths and limitations of this study:*

- The study will use a specifically pain oriented methodological approach that intends to go beyond purely informational pain neuroscience education, aiming for a more useful understanding of pain from patients.
- Incorporating the salutogenic model in therapeutic education will bring a health focused perspective to the research, potentially enriching its interpretive framework helping future interventions.
- The proposed use of clear and replicable guidelines such as GREET or TIDieR may help in the practical implementation challenge for clinicians, potentially influencing also the study’s reproducibility.
- While the study plans to assess psychosocial factors, pain intensity levels, and brain plastic changes due to therapeutic education, this comprehensive approach might pose notable methodological challenges.
- Given the emerging field of brain plasticity related to therapeutic education in chronic pain patients, the limited existing research may influence the interpretive depth of the study’s findings.

*Protocol version:* In this first version of the protocol dated July 2023, we are recruiting the sample, which is planned to be completed in approximately two months (September 2023) in order to start the trial in October 2023.

## Background

Chronic pain is a prevalent health problem that affects one in five people worldwide [1]. It is a debilitating condition that can have a significant impact on a person’s quality of life, affecting their ability to perform daily activities and decreasing their emotional and physical well-being [2], as well as the resources needed to cope [3]. It is crucial to keep in mind one serious drawback that accompanies chronic pain, the frequent dependence on drugs for pain relief. In particular, the use of opioids has become one of the main medications for treating pain, but at the same time, one of the causes of the increase in mortality due to overdose [4] . However, physiotherapy modalities that include therapeutic education and exercise are a common non-pharmacological therapeutic approach for chronic pain and its maladaptive plasticity [6]–[8], defined as a change in the structure or function of the nervous system that results in increased pain sensitization [9], [10]. Specifically, therapeutic education (TE) plays a particularly important role in reversing maladaptive plasticity, reducing pain, disability, fear avoidance, and pain catastrophizing, among other aspects [11] TE is a fundamental component in the management of chronic pain and addresses a variety of topics that are essential for improving patients’ quality of life [12], [13]. These topics include understanding pain and its mechanism of production [14], promoting self-regulation and self-care strategies [15], acquiring coping skills [16], reducing fear-avoidance and pain catastrophizing [17], as well as promoting healthy lifestyles and optimizing physical functionality[18]. One of the main goals for the patient is to reconfigure maladaptive beliefs associated with their chronic pain, involving a change in the structure and function of the nervous system that has contributed to increased pain sensitization [19], [20].

However, not any type of educational approach is sufficient to achieve this. There are many types of therapeutic education, addressing different topics [21], [22] and using different dosages [23]–[25], which inevitably lead to different results [26]–[28]. A specific approach is required that considers both the biological and behavioral aspects of chronic pain. Furthermore, in order to accurately determine the true impact of this type of non-only-informative intervention, it is imperative to measure and evaluate the effects of the TE not only in terms of perceived pain and psychosocial factors, but also through the lens of brain plasticity biomarkers and draw correlations within these domains.

In this sense, it is essential to consider biological components such as brain-derived neurotrophic factor (BDNF) and its relationship to cerebral plasticity in patients with persistent pain [29], [30]. BDNF, a protein that plays an important role in pain sensitization and neuronal plasticity, has been associated with neuronal hyperexcitability in chronic pain [31]. Understanding its relationship to pain is crucial for developing new therapies. Manipulating BDNF expression could offer options for modulating neuronal plasticity and relieving pain in patients with chronic pain. However, the impact of these interventions on the adaptive changes in the brain, which are crucial in patients with persistent pain, remains largely unknown. Additionally, the lack of adherence to established guidelines for reporting interventions, such as the TIDieR checklist for general interventions [32], or GREET checklist for educational interventions [33] and the CERT checklist for exercise interventions [34]hinders their replication and adaptation to individual patients, thus limiting their practical utility [35].

In summary, despite the challenges that exist in the field of education and exercise for chronic pain management, this model of pain therapeutic education will address the mentioned needs, providing a comprehensive and evidence-based approach that considers changes in brain plasticity learning related, biobehavioral aspects, appropriate dosing, and objective evaluation of results.

## Materials and Methods

### Study Setting

This proposed study is a single-blind, parallel-group, randomized control trial. This study protocol follows the structure based on the PICO question (Population, Intervention, Comparison, Outcome) [36] and adheres to the recommendations of the Standard Protocol Items: Recommendations for Interventional Trials (SPIRIT) guideline [37]. In addition, specific interventions were described based on the Template for Intervention Description and Replication (TIDieR) checklist [32], the Guideline for Reporting Evidence-based practice Educational interventions and Teaching (GREET) [33], and the Consensus on Exercise Reporting Template (CERT) checklist [34]. The trial has been approved by the ethics committee of Rey Juan Carlos University (Ethical number: 1901202202822) and the entire study will be conducted at the Universidad Rey Juan Carlos located in Alcorcón, Madrid (Spain). The trial identifier code is NCT05623579, and it was registered on November 18, 2022, in ClinicalTrials.gov.

### Eligibility criteria

Patients will be recruited from the physical therapy clinics located in Madrid, Spain. The participants will be included according to the following inclusion criteria: aged between 18 and 65, having a history of pain lasting more than twelve weeks, pain of at least 3 out of 10 on the visual analogue scale, possessing the ability to comprehend, speak, and write in Spanish, not currently undergoing any other treatments up to 3 months prior to the study, and explicitly accepting that they will not be allowed to initiate new treatments until the end of the follow-up period. The exclusion criteria are neurological cognitive alteration that prevents understanding the contents of the educational program (in case of doubt, assessment with the Mini Mental test will be conducted with minimum score of 25 required), systemic, oncological or inflammatory diseases; psychiatric pathologies, pregnancy, diabetes type II and the lack of ability to adhere to protocol.

The following professionals were selected to carry out the study: two physiotherapists with more than 15 years of experience in the treatment of chronic pain for the pain education and exercise therapy component, two nurses for blood collection, another physiotherapist for administering questionnaires on psychosocial variables and quantitative sensory testing, and two biochemists for the analysis of BDNF.

### Interventions

All patients who meet the inclusion criteria and agree to participate in the study will be informed verbally and in writing with the delivery of the information sheet of the specific characteristics of the study. They will be asked to sign the written informed consent form to agree to participate.

Sociodemographic data will then be collected and psychosocial variables on their current pain process. This data collection will be carried out in a room located in the physiotherapy gymnasium of the university center, previously designated for measurements by the evaluating researcher as well as blood extraction.

Once the specific questionnaires and the pressure pain threshold (PPT) at the joints of the elbow and tibialis anterior are measured, a trained professional will collect 5 ml of blood from the ulnar vein of the participants in a citrate vacutainer. The collection of the blood sample will always take place between 6 and 7 p.m. to minimize the possible effects of circadian changes [38]–[40]. Blood samples will be collected in tubes containing K3-EDTA and centrifuged at 1000 g for 10 min at 4 °C. The supernatants for the total plasma (TP) condition shall be frozen at −80°C. Blood collection will be performed in both groups at rest before the intervention, 15 days after the intervention, at the end of the intervention period, 1 month and 8 months after the intervention.

After the initial data collection, a researcher other than the initial evaluator, and the physiotherapist-therapist who will apply the subsequent treatment to ensure blinding, will proceed to randomly assign the subjects to one of the two study branches by means of a list previously generated with GraphPad software (version 5.01 Graphpad software, Inc. San Diego, California, USA).

### Intervention group (Exercise + POBTE)

Patients assigned to the intervention group will be instructed by a physiotherapist in the performance of the therapeutic exercise sessions according to their functional capacity, taking as a reference their HRmax [41]. They will be prescribed the reference intensity measures for the expected increase in training time and indications on potential risks, and references for the interruption of the exercise in their case. The total duration of the exercise program will be 4 weeks, with 3 sessions of a maximum of 45 minutes per session. On alternate days, twice a week the patient will receive *Pain Oriented Biobehavioral Therapeutic Education (POBTE)* sessions, focusing on the specific parameters described below and. The duration of the sessions will be a maximum of 45 minutes each day [42], and the education program will be 8 sessions, over 4 weeks. The intervention group will be instructed to refrain from discussing treatment details with participants assigned to the control group until the end of the treatment and follow up period.

The main learning areas that will be addressed and the biobehavioral interventions proposed are summarized in Figure S2, and all the conceptual framework of the POBTE intervention is explained in more detail in Annex S4. The specific cognitive components of the intervention are listed in the general intervention checklist, following the TiDier checklist (see Annex S1), also the exercise intervention is presented in Annex S2 with the CERT checklist, and the Therapeutic Educational Intervention is presented in Annex S3 with the GREET checklist.

A chronological representation of the tasks to be performed during the POBTE intervention is presented in Figure S3. This figure is arranged to correspond to the stages of the meaningful learning phase within the intervention design.

### Active Control group (Exercise)

Patients assigned to the active control group will be instructed by a physiotherapist in the performance of therapeutic exercise sessions according to their functional capacity, taking as a reference their HRmax. They will be prescribed the reference intensity measures for the expected increase in training time and indications on potential risks, and references for the interruption of the exercise in their case. The total duration of the exercise program will be 4 weeks, with 3 sessions of a maximum of 45 minutes per session. Contents of the exercise intervention are described in the CERT checklist (Annex S2).

### Interventions modifications

Criteria for discontinuing or modifying allocated interventions for a given trial participant may include participant request, adverse events or harms, improved or worsened disease, and non compliance. In the context of this study, criteria will be adapted to the specific needs and characteristics of the study population.

### Interventions adherence

To improve adherence, we will provide clear instructions, personalized feedback and support, simplify the intervention as much as possible without losing scientific rigor, and monitor attendance, self-reports, and reminders. Written materials and videos will be provided, along with personalized feedback, progress reports, and regular check-ins. The attendance, self reports and reminders will be used to monitor adherence.

### Outcomes

#### Primary outcome

##### Pain intensity

*Visual Analogue Scale (VAS):* Pain intensity will be measured using a 100 mm visual analogue scale where 0 represented ‘no pain’ and 100 the ‘worst pain imaginable’. Participants draw a mark at a point on the line that best reflects the pain they are experiencing at the time of measurement. Higher scores indicate higher pain levels. The sensitivity and specificity of this questionnaire and the acceptability of its psychometric properties have been previously approved [43].

#### Secondary outcomes

##### Biochemical

*Plasma BDNF levels:* Within 30 minutes after collection, blood samples shall be centrifuged and the plasma separated into 0.5 ml aliquots for further analysis. Plasma BDNF levels shall be determined by enzyme-linked immunosorbent assay (ELISA) test using monoclonal antibodies specific for neurotrophin (R&D Systems, Minneapolis, MN) using the manufacturer’s protocol. All samples shall be tested in duplicate to avoid intra-assay variations. The lower limit of detection for BDNF from the kit is 7.8 pg/mL. Assay (ELISA) using a ChemiKine BDNF ELISA kit, CYT306 (Chemicon/Millipore, Billerica, MA). Optical density will be measured using an ELISA reader at a wavelength of 450 nm (GloMax-Multi Microplate Reader, Promega, Madison, WI) for multiplex assay measurements. Data will be expressed in pg/mg protein.

##### Psychosocial

*Anxiety and depression:* will be assessed using the Spanish validated version of the HADS self-completion scale. This scale is divided into two subscales of 7 items each: 1) depression (HADSDep); and 2) anxiety (HADSAnx). The HADS has demonstrated good reliability and validity in various pathologies [45].

*Quality of life:* measured with the EuroQoL-5D questionnaire (EQ-5D), it is a self report instrument to assess health-related quality of life composed of three items, a 5-factor descriptive scale, a second item composed of a vertical VAS and a social values index generated by the instrument. The EQ-5D has shown good psychometric characteristics [46].

*Catastrophism*: will be measured with the Spanish version of the Pain Catastrophism Scale (ECD). This has demonstrated adequate psychometric characteristics for the assessment of this construct with internal validity (Cronbach’s alpha 0.81) [47].

*Chronic Pain Grading Scale (CPGS):* is a self-report instrument consisting of an 8-item scale. It has a high internal consistency, Cronbach’s α is 0.87, similar to that of other language versions, and the intraclass correlation coefficient is 0.81. The average administration time is 2 min 28 s [48].

*Tampa Scale for Kinesiophobia (TSK):* is a self-report questionnaire that consists of 17 items. The internal consistency of the TSK has been reported to be high, with Cronbach’s alpha coefficients ranging from 0.74 to 0.93, indicating strong reliability. Test-retest reliability has also been reported to be good, with correlation coefficients ranging from 0.75 to 0.88 [49].

*Pittsburgh Sleep Quality Index (PSQI):* is a self-rated questionnaire that consists of 19 items that are categorized into seven components, including subjective sleep quality, sleep latency, sleep duration, habitual sleep efficiency, sleep disturbances, use of sleep medication, and daytime dysfunction. The internal consistency of the PSQI has been reported to be high, with Cronbach’s alpha coefficients ranging from 0.77 to 0.83, indicating strong reliability. Test retest reliability has also been reported to be good, with correlation coefficients ranging from 0.85 to 0.87 [50].

*Perceived Stress Scale:* is a self-report questionnaire that consists of 10 items, each rated on a 5-point Likert scale ranging from 0 (never) to 4 (very often). The internal consistency of the PSS has been reported to be high, with Cronbach’s alpha coefficients ranging from 0.78 to 0.91, indicating strong reliability. Test-retest reliability has also been reported to be good, with correlation coefficients ranging from 0.67 to 0.85 [51].

*International Physical Activity Questionnaire (IPAQ):* is a self-reported questionnaire that consists of 27 items that assess the frequency, duration, and intensity of physical activity. The internal consistency of the IPAQ has been reported to be moderate to high, with Cronbach’s alpha coefficients ranging from 0.73 to 0.95. Test-retest reliability has also been reported to be good, with correlation coefficients ranging from 0.70 to 0.88 [52].

*Chronic Pain Self-Efficacy Scale (CPSS):* is a self-reported questionnaire that consists of 22 items that are rated on a 0-10 scale, with higher scores indicating greater self-efficacy in managing pain. The internal consistency of the CPSS has been reported to be high, with Cronbach’s alpha coefficients ranging from 0.89 to 0.95, indicating strong reliability. Test retest reliability has also been reported to be good, with correlation coefficients ranging from 0.63 to 0.90 [53].

*Knowledge questionnaire on specific aspects of pain:* ad hoc, which includes 5 questions in multi-choice test format on the contents used in the education sessions.

*Global Perception of Change:* the only one that will be tested after the intervention. This instrument consists of a 100mm line numbered in centimeters from -5 to 5 where the left end reads “Much Worse”, 0 is printed as “no change” and 5 is marked as “fully recovered”. On these values the patient is asked to answer the following question: “In relation to your pain, how would you describe your current state of health compared to when your pain began?” This version of the change perception scale has been assessed in the literature as the most appropriate in musculoskeletal pain processes, and as a valid and reliable instrument [54].

##### Others

*Pressure Pain Thresholds (PPT):* Pressure pain thresholds will be assessed using a pressure algometer. The algometer will be applied to different points in the body (elbow and knee) and participants will be asked to indicate the point at which pressure changes to pain. The pressure will be gradually increased at a rate of approximately 1 kg/s until the participant indicates that they feel pain. The threshold pressure at which the sensation changes from pressure to pain will be recorded in kilograms (kg) and will be used as an indicator of pressure pain threshold. Higher PPT values indicate higher pain thresholds. The validity and reliability of using a pressure algometer to measure PPT have been previously established [44].

Height and Weight: both variables will be determined by patient estimation, with no exploration of these variables in the measurement process.

##### Sociodemographic

Age, Marital status (Single, Married, Divorced, Widowed), Employment status (Active, Unemployed, Retired, Temporary disability, Student, Homemaker, Other), Educational level (none, primary, secondary, university), Gender.

##### Exercise

Among the exercise variables, the parameters described below will be considered:

*Blood pressure and respiratory rate:* Measured with the Anura application, which has demonstrated a reliability with conventional blood pressure measurement of 98% [55].

*Saturation and HR:* Measured with an Oxylink finger pulse oximeter.

All the outcome measurements will be assessed in a random order at baseline, at the halfway point of the treatment, post intervention, and one- and eight-months post intervention. (Figure S4, S5 and S1).

### Participants timeline

We have included a schematic diagram outlining the time schedule of participant enrolment, interventions, assessments, and visits, as shown in the following figures (S4 and S5).

### Sample Size

The sample size was calculated using a t-test design for independent means, using the G*Power program (Universität Düsseldorf, Germany) and establishing a significance level of 0.05 and a power of 90%.

To determine the effect size, Cohen’s d formula was used, which compares the mean difference on the Visual Analogue Scale (VAS), set at 2 points, and the pooled standard deviation (SDpooled). Based on data from a previous pilot study, the standard deviations for the intervention and control groups were 1.29 and 2.61 respectively. Using these data, the SDpooled was calculated using the formula for independent groups: sqrt[(((n1-1)*SD1^2 + (n2-1)SD21^2] / (n1+n2-2)], yielding a value of 2.13 [56].

Therefore, the effect size (d) was calculated as 2/2.13 = 0.94. With an allocation ratio of 1:1 between groups, these data were entered into GPower [57], resulting in the need for at least 25 subjects per group to be able to detect a 2-point difference in VAS with 90% power.

Considering a loss rate of 25%, the final sample size was adjusted to 32 subjects per group. Consequently, the study will require a total of 64 subjects to reach the required statistical power.

### Recruitment

To achieve our target sample size, from July to September 2023, we will partner with physiotherapy clinics, distribute pamphlets, utilize social media, recruit through support groups and emphasize the benefits of the intervention.

### Allocation

#### Sequence generation, concealment mechanism and implementation

Individuals who met the inclusion criteria will randomly be allocated to one of two treatment arms: (1) POBTE plus exercise or (2) exercise therapy, using computer-generated random numbers in stratified permuted block (block size of 4 and 6). The allocation will be concealed in an opaque, sealed envelope. A research assistant opens them and assigns patients to either of the treatment arms. The randomization will be conducted after signing the informed consent and baseline assessments.

### Blinding

#### Masking

In this trial, outcome assessors and data analysts will be blinded after intervention assignment. They won’t know the participants’ intervention group during outcome assessment or data analysis. Due to the intervention’s nature, involving educational sessions and exercises requiring interaction, participants and educators cannot be blinded.

#### Emergency unmasking

A secure process, such as a password-protected system, will be employed. Unblinding should only occur when necessary, preserving the trial’s integrity.

### Data Collection plan

The trial will collect baseline and outcome data through questionnaires, laboratory tests, and physical assessments, with standardized and validated instruments to ensure data quality. The instruments used in the trial are validated, and information on them can be found in the protocol.

### Data collection plan: retention

To ensure that participants remain engaged and complete the follow-up in our trial, we have implemented several strategies. Firstly, we provide clear and concise information to our participants regarding the trial, including potential benefits and risks, to help them understand the importance of their participation. Secondly, we establish clear communication channels between the research team and participants to encourage an open and trusting relationship, where participants feel comfortable asking questions and providing feedback. Additionally, we make sure to conduct regular follow-up phone calls or emails to remind participants of upcoming study visits and address any concerns they may have.

### Data Management

All data will be entered into a secure database system with restricted access, and data quality will be maintained with range checks and double data entry. Data security and storage plans include regular backups, restricted access, and secure transfer methods.

### Statistics: outcome

Data analysis will be performed with SPSS statistical software version 25.0 (SPSS Inc., Chicago, IL, USA). A descriptive analysis of the demographic characteristics and pain intensity of the sample will be performed, presenting continuous variables as Mean ± Standard Deviation (SD), 95% Confidence Interval (CI), while categorical variables will be presented as Number (n) and Percentage (relative frequency, %). If the quantitative variables follow a non-normal distribution, they will be described with the median and the interquartile range.

Parametric tests (normal distribution) will be chosen for the comparison between groups based on the central limit theorem, since the sample size of both groups will be greater than 30. Therefore, the one-way ANOVA test will be used to analyze the group factor in the quantitative variables and the chi-square test for the qualitative variables. If the ANOVA reveals statistical significance, a post-hoc analysis with Bonferroni correction will be performed. A P-value < 0.05 will be accepted as statistically significant.

### Statistics: additional analyses

We will conduct secondary analyses to examine treatment effects within specific subgroups of participants and adjust for potential confounding variables. Subgroup analyses will consider age, gender, and duration of pain. Multiple linear regression models will be used for adjusted analyses, and sensitivity analyses will be conducted to assess the robustness of our results.

### Statistics: Analyses population and missing data

The analysis will be based on the intention-to-treat principle, including all randomized participants in the analysis, regardless of their compliance with the intervention protocol. Missing data will be handled using appropriate imputation methods, such as multiple imputations or last observation carried forward. Sensitivity analyses will be conducted to evaluate the impact of different imputation methods on the results. The analysis population will be clearly defined and any deviations from the intention-to-treat principle will be reported and justified.

### Data monitoring: Formal committee

This small-scale, low-risk intervention trial does not require a Data Monitoring Committee (DMC). Instead, the study investigators will take on the responsibility of monitoring trial data. The primary investigator will regularly review study data to ensure adherence to the protocol and to maintain the safety and well-being of participants. Should significant safety concerns or other issues arise, the primary investigator will collaborate with the study team and, if necessary, consult with an independent expert to determine the best course of action. Any concerns or protocol deviations will be reported to relevant regulatory authorities promptly.

### Data monitoring: Interim analysis

Although data and blood samples will be collected at 15 days during the intervention period, no interim analyses are planned for this trial, although it might be interesting to see if there are possible changes in BDNF levels, as a biomarker of brain plasticity, in relation to the learning curve [58] experienced by the patients. It is expected that the learning process they experience during the intervention will lead to significant changes in BDNF levels.

### Harms

The research team will monitor participants for adverse events related to the intervention or trial conduct. Solicited events will be assessed during follow-up visits, while spontaneously reported events will be identified during unscheduled interactions.

Adverse events will be documented, and the principal investigator will review them, determining if they require reporting to the ethics committee and regulatory authorities. Unintended effects of the trial interventions or conduct will also be monitored and managed.

All events will be reported in the trial report, publication, and to the ethical committee.

### Research ethics approval

To get approval for our study from a research ethics committee (REC), we developed a detailed protocol, submitted necessary documentation, addressed any questions or concerns from the REC, obtained REC approval and will maintain ongoing compliance. We also have plans to communicate important protocol modifications to investigators, the REC, trial participants, trial registries, journals, and regulators.

### Consent or assent

Informed consent or assent will be obtained from potential trial participants or authorized surrogates by trained research staff. The consent process will involve the use of written informed consent documents, which will be provided to the potential participants or their authorized surrogates in advance of the trial. The documents will include a description of the study, the risks and benefits of participation, and the rights and responsibilities of the participants. The research staff will also be available to answer any questions or concerns that may arise during the consent process.

### Consent or assent: ancillary studies

We included in the informed consent form for potential ancillary studies involving participant data and biological specimens. The form will state that data and specimens may be used for future research with ethical approval, and participants could withdraw consent at any time. This ensured participants were fully informed and their rights and privacy were protected.

### Confidentiality

We will protect participants’ personal information throughout the trial. During screening, we will only collect necessary information, and destroy any unused data. Personal information will be kept secure and restricted to authorized members of the study team during the trial, identified only by participant ID numbers. After the trial, identifying information will be removed, and data will be stored securely for a specified period. We will inform participants of our confidentiality policies during informed consent.

### Data access

Access to the final trial dataset will be limited to authorized members of the research team, including the principal investigators and their designated staff.

### Ancillary and post-trial care

The study team will develop a plan for providing ancillary or post-trial care for harm to participants during a clinical trial. They will inform participants about these plans during the informed consent process and work with regulatory authorities and review boards to ensure appropriate provisions are in place.

### Dissemination policy: Trial results

We will communicate trial results to participants, healthcare professionals, and the public. Results will be published in a peer-reviewed journal, presented at conferences, and reported in publicly accessible databases. We’ll provide lay summaries to participants and ensure healthcare professionals receive information. We’ll publish regardless of the outcome, share the protocol, dataset and statistical analysis, and uphold responsible research guidelines. Our communication will be clear, timely, and accurate with integrity and transparency.

### Dissemination policy: Authorship

Authorship eligibility guidelines will follow international standards [59]. Contributions of eligible individuals will be clearly stated in publications, while professional writers will only be acknowledged if they meet the authorship eligibility guidelines.

### Dissemination policy: Reproducible research

After the study, we will share the full protocol, de-identified participant-level dataset, and statistical analysis to encourage transparency and enable further research. We will provide technical support and educational resources to promote accessibility and reproducibility of study findings.

## Discussion

The current randomized controlled trial (RCT) seeks to examine the impact of a specific therapeutic education program, combined with exercise, on BDNF levels and pain perceived intensity among individuals suffering from chronic musculoskeletal pain. This article not only aims to offer a more rigorous approach to therapeutic education from a biobehavioral perspective, but also assesses the outcomes of an intervention that encompasses both the cognitive aspect and pain-related plasticity biomarkers, such as BDNF. Consequently, it represents a valuable contribution to low-cost, non-pharmacological treatments in physiotherapy. Although the study design and methods have been meticulously planned, there are several practical and operational considerations that need to be addressed.

One potential limitation of our study relates to the heterogeneity of chronic pain. Despite having well-defined inclusion criteria, it is important to recognize that chronic pain is a complex and multifactorial condition [60]–[62]. While we have not imposed restrictions on the specific site of pain in our selection criteria, supported by evidence that chronic pain is centrally mediated rather than solely dependent on the location of the injury [63]–[66], it is evident that various individual factors influence the pain experience in each patient [67]–[69], which may impact the obtained results. To address this limitation, in addition to the inclusion criteria, we will consider conducting subgroup analysis if necessary and feasible. This analysis will allow us to examine possible clusters of patients with similar characteristics, which will enhance our understanding of differences and treatment responses within our sample.

Another concern is the possible confounding effects of medications and other interventions that participants might be undergoing. To address this issue, participants will be strictly advised to continue their regular medication regimen (if applicable) but will not be permitted to initiate any new treatments during the program and follow-up period. This measure is implemented to minimize potential confounding factors and ensure the integrity of the study results. The study team will closely screen and monitor participants to ensure compliance with this requirement.

Additionally, statistical analysis will consider adjusting for any confounding variables that may arise.

Finally, it is important to mention that the measurement of BDNF levels can pose challenges due to the inherent variability in sample collection, storage, and analysis. In this study, BDNF levels will be specifically measured in plasma samples, as they have been found to be more stable during the analysis compared to serum samples [70]. To ensure precise and reliable results, the study team will adhere to rigorous protocols for plasma sample collection, processing, and storage. The analysis will be conducted using standardized ELISA kits with regular calibration and quality control checks implemented to minimize potential sources of variation. These measures are crucial for ensuring the accuracy and validity of the BDNF measurements and upholding the integrity and reliability of the study’s findings.

In conclusion, this randomized controlled trial (RCT) serves as a significant milestone in two important aspects. Firstly, it contributes to the development of a more structured and rigorous therapeutic education model for individuals with chronic musculoskeletal pain by implementing validated checklists and protocols [32], [33]. This standardized approach ensures the delivery of effective interventions tailored to the specific needs of these patients.

Secondly, the study investigates the changes occurring at the central nervous system level by measuring biomarkers such as BDNF. This provides valuable insights into the underlying mechanisms of chronic pain and the impact of non-pharmacological interventions on brain plasticity. By elucidating the relationship between therapeutic education, BDNF levels, and pain outcomes, this research can pave the way for novel and targeted strategies to enhance treatment efficacy and improve the quality of life for individuals suffering from chronic pain.

## Data Availability

No datasets were generated or analysed during the current study. All relevant data from this study will be made available upon study completion

## Appendices

### Informed consent materials

We will attach the model consent form and related documentation to the annexes section of our study to ensure a clear record of the consent process that can be easily accessed and verified by future reviewers.

### Biological specimens

We will collect and label blood samples, isolate plasma, and measure BDNF levels using an ELISA kit. Quality control measures will be in place to ensure accurate measurements. Specimens will be stored in secure, monitored freezers (−80°) at the research site and laboratory, and tracked using unique participant identifiers. With proper ethics approval and participant consent, stored specimens may be used in future ancillary studies. This plan ensures the proper handling and availability of biological specimens for future research.

## Acknowledgments

This study is part of PhD thesis of Silvia Di Bonaventura and will be supported by Spanish Ministry of Education (Grant number: FPU20/00041).

## List of acronyms

BDNF: Brain Derived Neurotrophic Factor
CERT: Consensus on Exercise Reporting Template
CP: Chronic Pain
CPGS: Chronic Pain Grading Scale
CPSS: Chronic Pain Self-Efficacy Scale
ECD: Pain catastrophizing scale
EQ-5D: EuroQol-5 Dimensions
GREET: Guideline for Reporting Evidence-based practice Educational interventions and Teaching
HADS: Hospital Anxiety and Depression Scale
HR: Heart Rate
IPAQ: International Physical Activity Questionnaire
PICO: Population, Intervention, Control, Outcomes
PPT: Pressure Pain Threshold
POBTE: Pain Oriented Biobehavioral Therapeutic Education
PSQI: Pittsburgh Sleep Quality Index
PSS: Perceived Stress Scale
REC: Research Ethic Committee
RCT: Randomized Controlled Trial
SPIRIT: Standard Protocol Items: Recommendations for Interventional Trial
TE: Therapeutic Education
TIDieR: Template for Intervention Description and Replication
TP: Total Plasma
TSK: Tampa Scale for Kinesiophobia
VAS: Visual Analogue Scale

---

**Fig S1.**
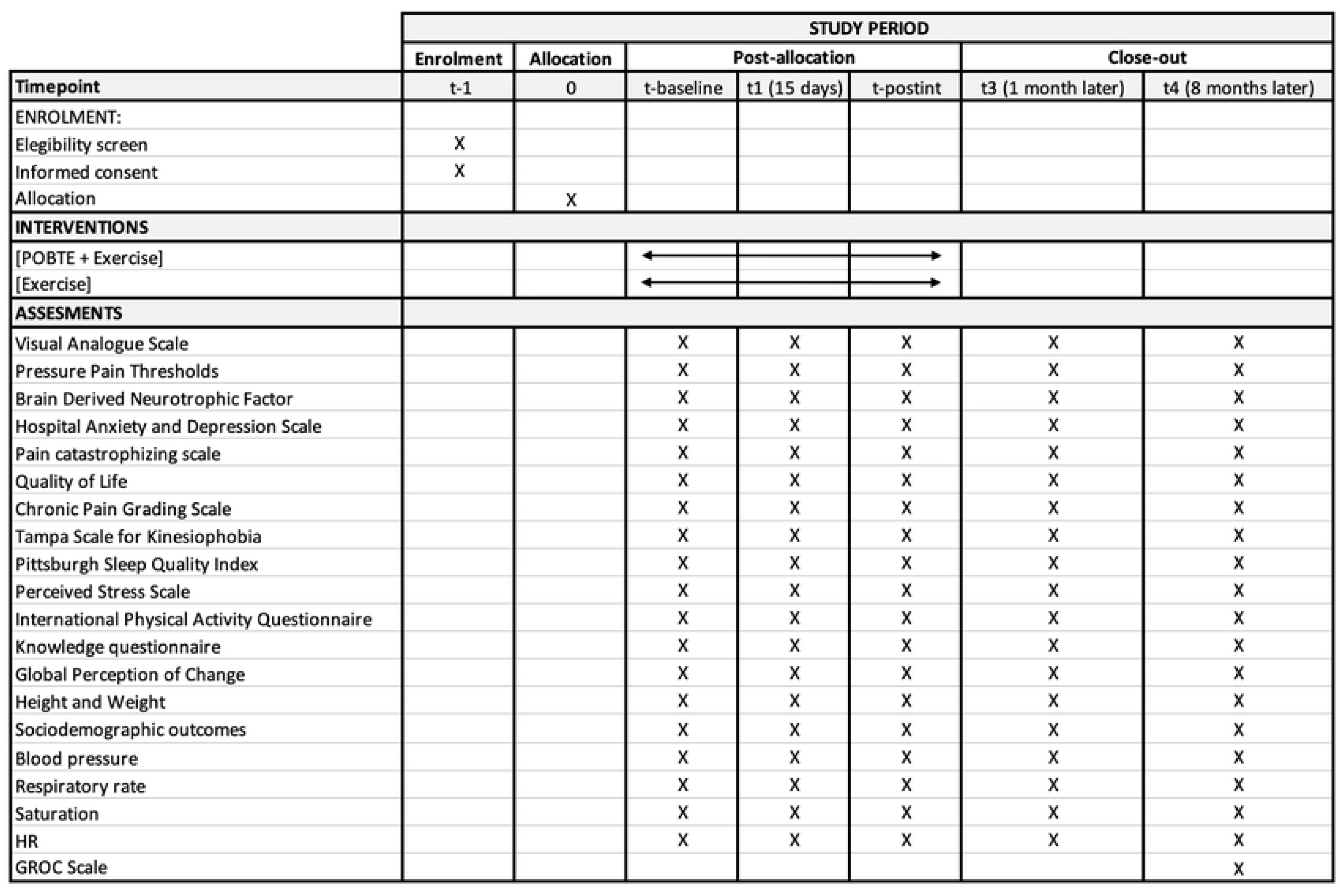
SPIRIT figure showing the schedule of enrolment, interventions, and assessments.

**Fig. S2.**
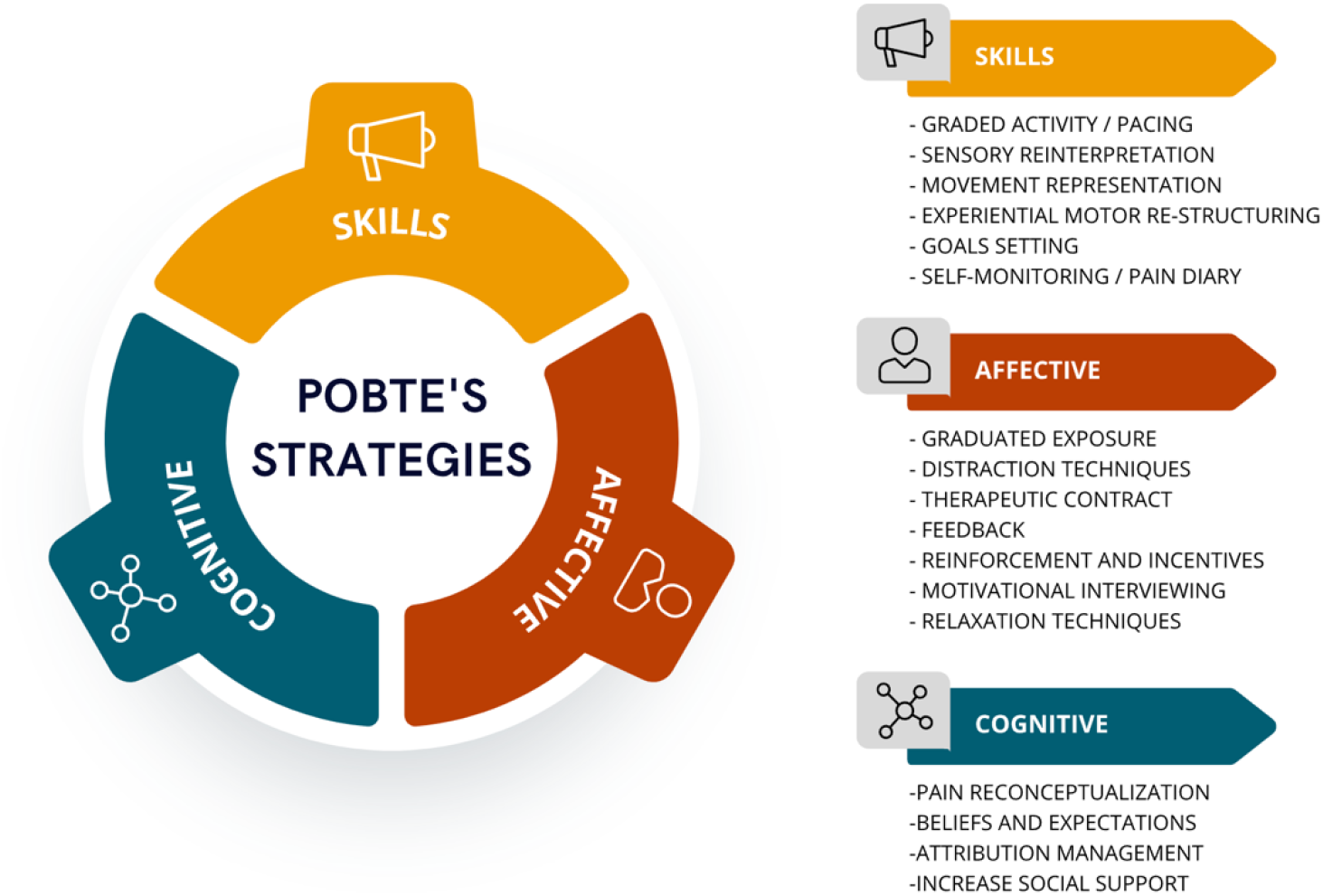
POBTE’s strategies.

**Fig. S3.**
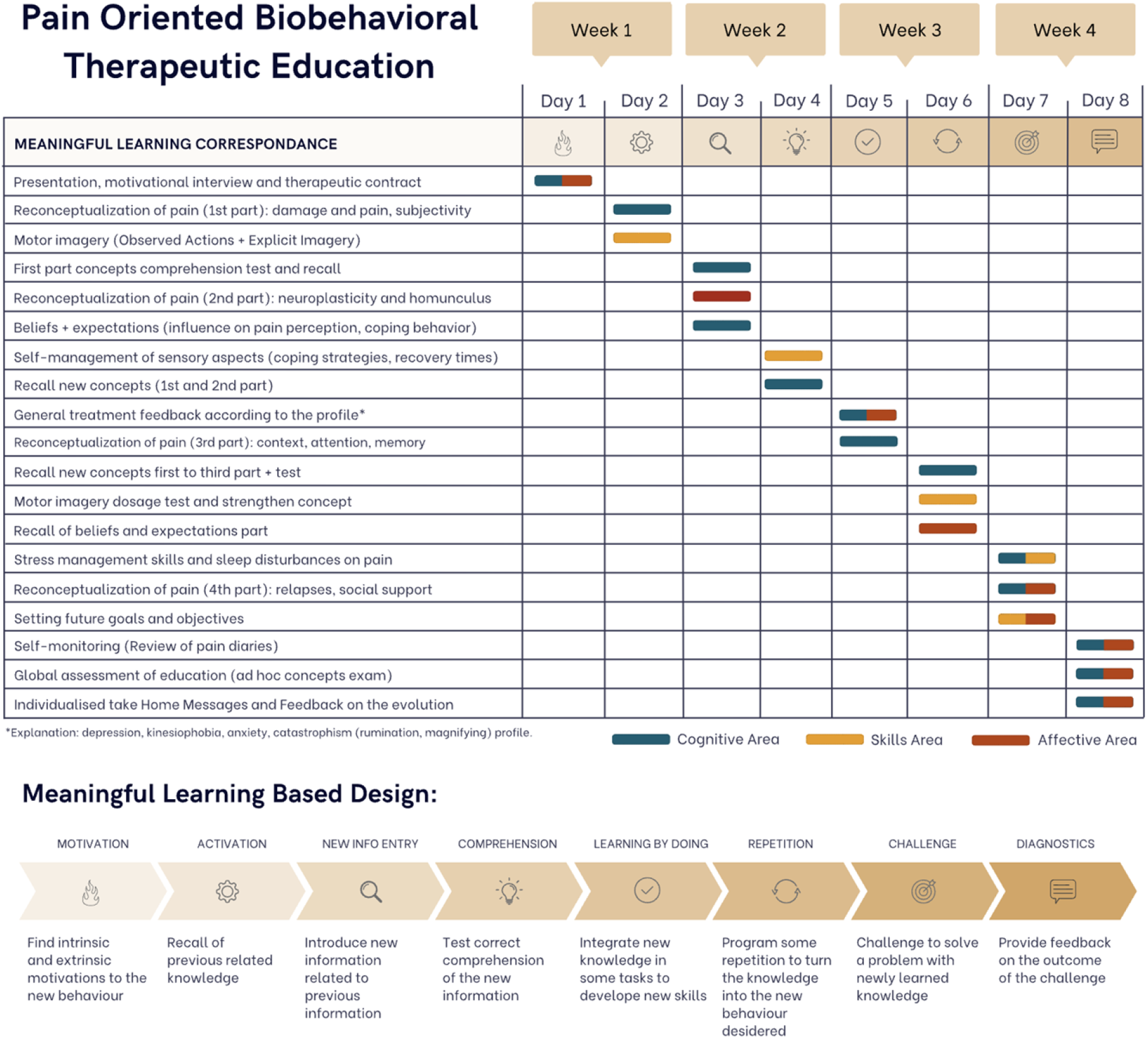
POBTE’s intervention.

**Fig S4.**
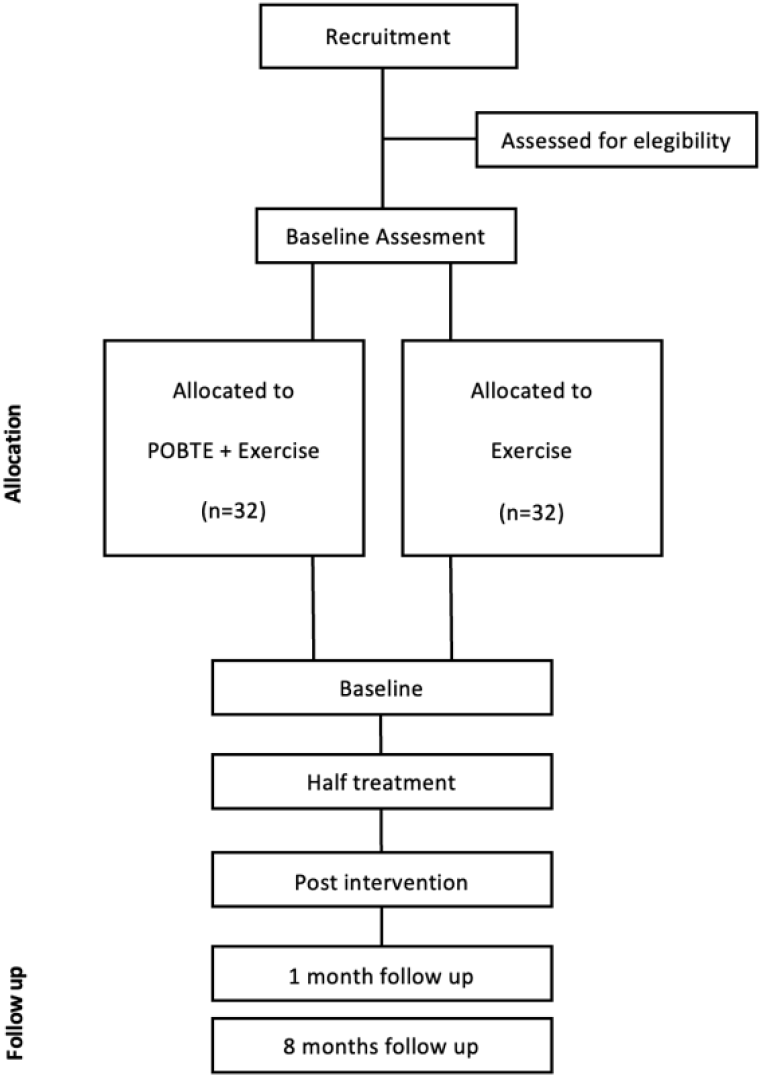
Flow diagram

**Fig S5.**
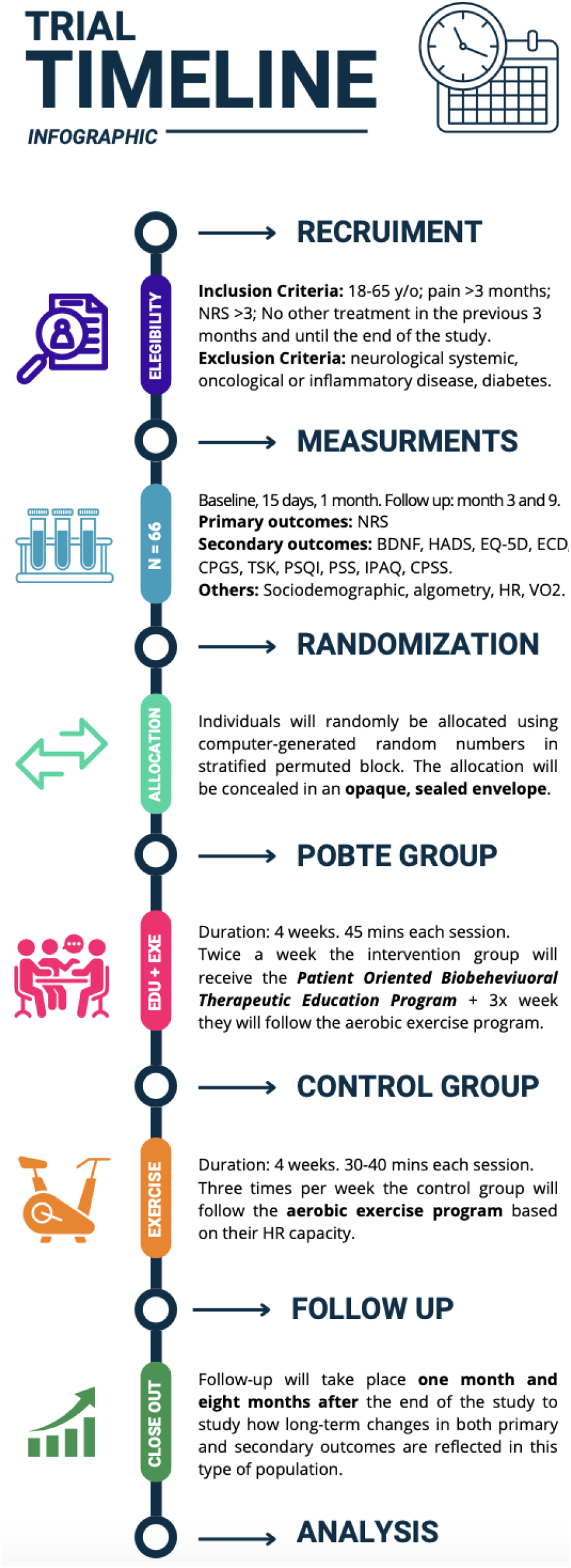
Trial Timeline

## References

[1] R. D. Treede et al., “Chronic pain as a symptom or a disease: The IASP Classification of Chronic Pain for the International Classification of Diseases (ICD-11),” Pain, vol. 160, no. 1. Lippincott Williams and Wilkins, pp. 19–27, Jan. 01, 2019. doi: 10.1097/j.pain.0000000000001384.

[2] S. E. E. Mills, K. P. Nicolson, and B. H. Smith, “Chronic pain: a review of its epidemiology and associated factors in population-based studies,” Br J Anaesth, vol. 123, no. 2, pp. e273–e283, Aug. 2019, doi: 10.1016/j.bja.2019.03.023.

[3] A. Büssing, T. Ostermann, E. Am Neugebauer, and P. Heusser, “Adaptive coping strategies in patients with chronic pain conditions and their interpretation of disease,” 2010. [Online]. Available: http://www.biomedcentral.com/1471-2458/10/507

[4] W. Häuser, T. Schubert, T. Vogelmann, C. Maier, M. A. Fitzcharles, and T. Tölle, “All-cause mortality in patients with long-term opioid therapy compared with non opioid analgesics for chronic non-cancer pain: A database study,” BMC Med, vol. 18, no. 1, Jul. 2020, doi: 10.1186/s12916-020-01644-4.

[5] W. Häuser, T. Schubert, T. Vogelmann, C. Maier, M. A. Fitzcharles, and T. Tölle, “All-cause mortality in patients with long-term opioid therapy compared with non opioid analgesics for chronic non-cancer pain: A database study,” BMC Med, vol. 18, no. 1, Jul. 2020, doi: 10.1186/s12916-020-01644-4.

[6] S. J. Snodgrass, N. R. Heneghan, H. Tsao, P. T. Stanwell, D. A. Rivett, and P. M. Van Vliet, “Recognising neuroplasticity in musculoskeletal rehabilitation: A basis for greater collaboration between musculoskeletal and neurological physiotherapists,” Man Ther, vol. 19, no. 6, pp. 614–617, Dec. 2014, doi: 10.1016/j.math.2014.01.006.

[7] E. Dayan and L. G. Cohen, “Neuroplasticity subserving motor skill learning,” Neuron, vol. 72, no. 3. pp. 443–454, Nov. 03, 2011. doi: 10.1016/j.neuron.2011.10.008.

[8] J. El-Sayes, D. Harasym, C. V. Turco, M. B. Locke, and A. J. Nelson, “Exercise Induced Neuroplasticity: A Mechanistic Model and Prospects for Promoting Plasticity,” Neuroscientist, vol. 25, no. 1. SAGE Publications Inc., pp. 65–85, Feb. 01, 2019. doi: 10.1177/1073858418771538.

[9] C. J. Woolf, “Central sensitization: Implications for the diagnosis and treatment of pain,” Pain, vol. 152, no. SUPPL.3. Mar. 2011. doi: 10.1016/j.pain.2010.09.030.

[10] X. Y. Li, Y. Wan, S. J. Tang, Y. Guan, F. Wei, and D. Ma, “Maladaptive Plasticity and Neuropathic Pain,” Neural Plasticity, vol. 2016. Hindawi Publishing Corporation, 2016. doi: 10.1155/2016/4842159.

[11] A. Louw, J. Nijs, and E. J. Puentedura, “A clinical perspective on a pain neuroscience education approach to manual therapy,” Journal of Manual and Manipulative Therapy, vol. 25, no. 3, pp. 160–168, May 2017, doi: 10.1080/10669817.2017.1323699.

[12] I. Saracoglu, M. I. Arik, E. Afsar, and H. H. Gokpinar, “The short-term effects of neuroscience pain education on quality of life in patients with chronic low back pain: A single-blinded randomized controlled trial,” Eur J Integr Med, vol. 33, Jan. 2020, doi: 10.1016/j.eujim.2019.101046.

[13] I. Saracoglu, M. I. Arik, E. Afsar, and H. H. Gokpinar, “The effectiveness of pain neuroscience education combined with manual therapy and home exercise for chronic low back pain: A single-blind randomized controlled trial,” Physiother Theory Pract, vol. 38, no. 7, pp. 868–878, 2022, doi: 10.1080/09593985.2020.1809046.

[14] M. Meeus, J. Nijs, J. Van Oosterwijck, V. Van Alsenoy, and S. Truijen, “Pain physiology education improves pain beliefs in patients with chronic fatigue syndrome compared with pacing and self-management education: A double-blind randomized controlled trial,” Arch Phys Med Rehabil, vol. 91, no. 8, pp. 1153–1159, 2010, doi: 10.1016/j.apmr.2010.04.020.

[15] I. Aguirrezabal et al., “Effectiveness of a primary care-based group educational intervention in the management of patients with migraine: A randomized controlled trial,” Prim Health Care Res Dev, 2019, doi: 10.1017/S1463423619000720.

[16] A. Malfliet et al., “Blended-learning pain neuroscience education for people with chronic spinal pain: Randomized controlled multicenter trial,” Phys Ther, vol. 98, no. 5, pp. 357–368, May 2018, doi: 10.1093/ptj/pzx092.

[17] J. A. Watson et al., “Pain Neuroscience Education for Adults With Chronic Musculoskeletal Pain: A Mixed-Methods Systematic Review and Meta-Analysis,” Journal of Pain, vol. 20, no. 10. Churchill Livingstone Inc., pp. 1140.e1–1140.e22, Oct. 01, 2019. doi: 10.1016/j.jpain.2019.02.011.

[18] A. Louw, I. Diener, D. S. Butler, and E. J. Puentedura, “The effect of neuroscience education on pain, disability, anxiety, and stress in chronic musculoskeletal pain,” Archives of Physical Medicine and Rehabilitation, vol. 92, no. 12. pp. 2041–2056, Dec. 2011. doi: 10.1016/j.apmr.2011.07.198.

[19] A. May, “Chronic pain may change the structure of the brain,” Pain, vol. 137, no. 1. pp. 7–15, Jun. 30, 2008. doi: 10.1016/j.pain.2008.02.034.

[20] R. F. Smallwood et al., “Structural brain anomalies and chronic pain: A quantitative meta-analysis of gray matter volume,” Journal of Pain, vol. 14, no. 7. pp. 663–675, Jul. 2013. doi: 10.1016/j.jpain.2013.03.001.

[21] B. F. Dear et al., “The pain course: A randomised controlled trial comparing a remote delivered chronic pain management program when provided in online and workbook formats,” Pain, vol. 158, no. 7, pp. 1289–1301, Jul. 2017, doi: 10.1097/j.pain.0000000000000916.

[22] M. Mehlsen, L. Hegaard, E. Ørnbøl, J. S. Jensen, P. Fink, and L. Frostholm, “The effect of a lay-led, group-based self-management program for patients with chronic pain: A randomized controlled trial of the Danish version of the Chronic Pain Self Management Programme,” Pain, vol. 158, no. 8, pp. 1437–1445, Aug. 2017, doi: 10.1097/j.pain.0000000000000931.

[23] “darnall2021”.

[24] G. Lorimer Moseley, M. K. Nicholas, and P. W. Hodges, “A Randomized Controlled Trial of Intensive Neurophysiology Education in Chronic Low Back Pain.”

[25] R. B. Saper et al., “Yoga, physical therapy, or education for chronic low back pain: A randomized noninferiority trial,” Ann Intern Med, vol. 167, no. 2, pp. 85–94, Jul. 2017, doi: 10.7326/M16-2579.

[26] A. Louw, E. J. Puentedura, K. Zimney, and S. Schmidt, “Know Pain, know gain? A perspective on pain neuroscience education in physical therapy,” Journal of Orthopaedic and Sports Physical Therapy, vol. 46, no. 3. Movement Science Media, pp. 131–134, Mar. 01, 2016. doi: 10.2519/jospt.2016.0602.

[27] L. J. Geneen et al., “Effects of education to facilitate knowledge about chronic pain for adults: A systematic review with meta-analysis,” Syst Rev, vol. 4, no. 1, Oct. 2015, doi: 10.1186/s13643-015-0120-5.

[28] L. Wood and P. A. Hendrick, “A systematic review and meta-analysis of pain neuroscience education for chronic low back pain: Short-and long-term outcomes of pain and disability,” European Journal of Pain (United Kingdom), vol. 23, no. 2. Blackwell Publishing Ltd, pp. 234–249, Feb. 01, 2019. doi: 10.1002/ejp.1314.

[29] A. Merighi et al., “BDNF as a pain modulator,” Progress in Neurobiology, vol. 85, no. 3. pp. 297–317, Jul. 2008. doi: 10.1016/j.pneurobio.2008.04.004.

[30] T. Brigadski and V. Leßmann, “BDNF: a regulator of learning and memory processes with clinical potential,” eNeuroforum, vol. 5, no. 1, pp. 1–11, Mar. 2014, doi: 10.1007/s13295-014-0053-9.

[31] J. Nijs et al., “Brain-derived neurotrophic factor as a driving force behind neuroplasticity in neuropathic and central sensitization pain: A new therapeutic target?,” Expert Opinion on Therapeutic Targets, vol. 19, no. 4. Informa Healthcare, pp. 565–576, Apr. 01, 2015. doi: 10.1517/14728222.2014.994506.

[32] T. C. Hoffmann et al., “Better reporting of interventions: Template for intervention description and replication (TIDieR) checklist and guide,” BMJ (Online), vol. 348, Mar. 2014, doi: 10.1136/bmj.g1687.

[33] A. C. Phillips et al., “Development and validation of the guideline for reporting evidence-based practice educational interventions and teaching (GREET),” BMC Med Educ, vol. 16, no. 1, Sep. 2016, doi: 10.1186/s12909-016-0759-1.

[34] S. C. Slade et al., “Consensus on Exercise Reporting Template (CERT): Modified Delphi Study,” 2016. [Online]. Available: http://www.equator-

[35] E. K.-Y. H. P. H. F. C. G.-M. O. V.-A. and J. M.-S. Javier Martinez-Calderon, “A Call for Improving Research on Pain Neuroscience Education and Chronic Pain: An Overview of Systematic Reviews,” Journal of Orthopaedic & Sports Physical Therapy, vol. 53:6, pp. 353–368, 2023.

[36] S. Aslam and P. Emmanuel, “Formulating a researchable question: A critical step for facilitating good clinical research,” Indian Journal of Sexually Transmitted Diseases, vol. 31, no. 1. pp. 47–50, Jan. 01, 2010. doi: 10.4103/0253-7184.69003.

[37] A. W. Chan et al., “SPIRIT 2013 statement: Defining standard protocol items for clinical trials,” Annals of Internal Medicine, vol. 158, no. 3. American College of Physicians, pp. 200–207, Feb. 05, 2013. doi: 10.7326/0003-4819-158-3-201302050-00583.

[38] S. W. Cain et al., “Circadian rhythms in plasma brain-derived neurotrophic factor differ in men and women,” J Biol Rhythms, vol. 32, no. 1, pp. 75–82, Feb. 2017, doi: 10.1177/0748730417693124.

[39] S. W. Choi, S. Bhang, and J. H. Ahn, “Diurnal variation and gender differences of plasma brain-derived neurotrophic factor in healthy human subjects,” Psychiatry Res, vol. 186, no. 2–3, pp. 427–430, Apr. 2011, doi: 10.1016/j.psychres.2010.07.028.

[40] A. A. H. Ali and C. von Gall, “Adult Neurogenesis under Control of the Circadian System,” Cells, vol. 11, no. 5. MDPI, Mar. 01, 2022. doi: 10.3390/cells11050764.

[41] C. J. Keating et al., “Utilizing Age-Predicted Heart Rate Maximum to Prescribe a Minimally Invasive Cycle Ergometer HIIT Protocol in Older Adults: A Feasibility Study.” [Online]. Available: http://www.intjexersci.com

[42] J. J. Amer-Cuenca et al., “How much is needed? Comparison of the effectiveness of different pain education dosages in patients with fibromyalgia,” Pain Medicine (United States), vol. 21, no. 4, pp. 782–793, 2020, doi: 10.1093/PM/PNZ069.

[43] A. M. Boonstra, H. R. Schiphorst Preuper, M. F. Reneman, J. B. Posthumus, R. E. Stewart, and R. Friesland, “Reliability and validity of the visual analogue scale for disability in patients with chronic musculoskeletal pain,” Wolters Kluwer Health | Lippincott Williams & Wilkins, 2008.

[44] A. M. Kinser, W. A. Sands, and M. H. Stone, “RELIABILITY AND VALIDITY OF A PRESSURE ALGOMETER.” [Online]. Available: www.nsca-jscr.org

[45] M. Carmen Terol-Cantero, V. Cabrera-Perona, and M. Martín-Aragón, “Revisión de estudios de la Escala de Ansiedad y Depresión Hospitalaria (HAD) en muestras españolas1,” Anales de Psicologia, vol. 31, no. 2, pp. 494–503, 2015, doi: 10.6018/analesps.31.2.172701.

[46] J. M. Cabasés, “The EQ-5D as a measure of health outcomes,” Gac Sanit, vol. 29, no. 6, pp. 401–403, Nov. 2015, doi: 10.1016/j.gaceta.2015.08.007.

[47] “CATASTR”.

[48] R. Ferrer-Peña, A. Gil-Martínez, J. Pardo-Montero, V. Jiménez-Penick, T. Gallego-Izquierdo, and R. La Touche, “Adaptación y validación de la Escala de gradación del dolor crónico al español,” Reumatol Clin, vol. 12, no. 3, pp. 130–138, May 2016, doi: 10.1016/j.reuma.2015.07.004.

[49] S. R. Woby, N. K. Roach, M. Urmston, and P. J. Watson, “Psychometric properties of the TSK-11: A shortened version of the Tampa Scale for Kinesiophobia,” Pain, vol. 117, no. 1–2, pp. 137–144, Sep. 2005, doi: 10.1016/j.pain.2005.05.029.

[50] C. L. Larche, I. Plante, M. Roy, P. M. Ingelmo, and C. E. Ferland, “The Pittsburgh Sleep Quality Index: Reliability, Factor Structure, and Related Clinical Factors among Children, Adolescents, and Young Adults with Chronic Pain,” Sleep Disord, vol. 2021, pp. 1–8, Apr. 2021, doi: 10.1155/2021/5546484.

[51] E. H. Lee, “Review of the psychometric evidence of the perceived stress scale,” Asian Nursing Research, vol. 6, no. 4. pp. 121–127, Dec. 2012. doi: 10.1016/j.anr.2012.08.004.

[52] C. L. Craig et al., “International physical activity questionnaire: 12-Country reliability and validity,” Med Sci Sports Exerc, vol. 35, no. 8, pp. 1381–1395, Aug. 2003, doi: 10.1249/01.MSS.0000078924.61453.FB.

[53] S. T. Cheng et al., “Developing a short multidimensional measure of pain self efficacy: The chronic pain self-efficacy scale-short form,” Gerontologist, vol. 60, no. 3, pp. E127–E136, Apr. 2020, doi: 10.1093/geront/gnz041.

[54] “gLOBAL RAtINg OF ChANge sCALes: A RevIew OF stReNgths AND weAkNesses AND CONsIDeRAtIONs FOR DesIgN.”

[55] C. Pham et al., “Effectiveness of consumer-grade contactless vital signs monitors: a systematic review and meta-analysis,” Journal of Clinical Monitoring and Computing, vol. 36, no. 1. Springer Science and Business Media B.V., pp. 41–54, Feb. 01, 2022. doi: 10.1007/s10877-021-00734-9.

[56] J. H. N. Cohen, “Statistical power analysis for the behavioral sciences (2nd ed.). .,” *Lawrence Erlbaum Associates*, 1988.

[57] F., E. E., L. A.-G., & B. A. Faul, “G*Power 3: A flexible statistical power analysis program for the social, behavioral, and biomedical sciences.,” Behavior Research Methods, 39, 175–191..

[58] J. M. J. Murre and J. Dros, “Replication and analysis of Ebbinghaus’ forgetting curve,” PLoS One, vol. 10, no. 7, Jul. 2015, doi: 10.1371/journal.pone.0120644.

[59] I. Vartiovaara, “Guidelines on authorship. International committee of medical journal editors,” Br Med J (Clin Res Ed), vol. 291, no. 6497, p. 722, 1985, doi: 10.1136/bmj.291.6497.722.

[60] I. Kela et al., “Chronic Pain: A Complex Condition With a Multi-Tangential Approach,” Cureus, Nov. 2021, doi: 10.7759/cureus.19850.

[61] B. Sherriff, C. Clark, C. Killingback, and D. Newell, “Impact of contextual factors on patient outcomes following conservative low back pain treatment: systematic review,” Chiropractic and Manual Therapies, vol. 30, no. 1. BioMed Central Ltd, Dec. 01, 2022. doi: 10.1186/s12998-022-00430-8.

[62] G. Rossettini, E. Carlino, and M. Testa, “Clinical relevance of contextual factors as triggers of placebo and nocebo effects in musculoskeletal pain,” BMC Musculoskeletal Disorders, vol. 19, no. 1. BioMed Central Ltd., Jan. 22, 2018. doi: 10.1186/s12891-018-1943-8.

[63] M. M. Volcheck, S. M. Graham, K. C. Fleming, A. B. Mohabbat, and C. A. Luedtke, “Central sensitization, chronic pain, and other symptoms: Better understanding, better management,” Cleveland Clinic journal of medicine, vol. 90, no. 4. NLM (Medline), pp. 245–254, Apr. 03, 2023. doi: 10.3949/ccjm.90a.22019.

[64] M.-A. Fitzcharles, S. P. Cohen, D. J. Clauw, G. Littlejohn, C. Usui, and W. Häuser, “Chronic Pain 2 Nociplastic pain: towards an understanding of prevalent pain conditions,” 2021. [Online]. Available: www.thelancet.com

[65] A. Latremoliere and C. J. Woolf, “Central Sensitization: A Generator of Pain Hypersensitivity by Central Neural Plasticity,” Journal of Pain, vol. 10, no. 9. pp. 895–926, Sep. 2009. doi: 10.1016/j.jpain.2009.06.012.

[66] J. Nijs et al., “Central sensitisation in chronic pain conditions: latest discoveries and their potential for precision medicine,” The Lancet Rheumatology, vol. 3, no. 5. Lancet Publishing Group, pp. e383–e392, May 01, 2021. doi: 10.1016/S2665-9913(21)00032-1.

[67] S. J. Linton, W. S. Shaw, and W. S. Shaw, “Impact of Psychological Factors in the Experience of Pain,” 2011. [Online]. Available: https://academic.oup.com/ptj/article/91/5/700/2735743

[68] L. M. Tracy, “Psychosocial factors and their influence on the experience of pain,” Pain Rep, vol. 2, no. 4, Jul. 2017, doi: 10.1097/PR9.0000000000000602.

[69] S. Bustan et al., “Psychological, cognitive factors and contextual influences in pain and pain-related suffering as revealed by a combined qualitative and quantitative assessment approach,” PLoS One, vol. 13, no. 7, Jul. 2018, doi: 10.1371/journal.pone.0199814.

[70] A. Polacchini et al., “A method for reproducible measurements of serum BDNF: Comparison of the performance of six commercial assays,” Sci Rep, vol. 5, Dec. 2015, doi: 10.1038/srep17989.

[71] S. Keen, M. Lomeli-Rodriguez, and A. C. D. C. Williams, “Exploring how people with chronic pain understand their pain: A qualitative study,” Scand J Pain, vol. 21, no. 4, pp. 743–753, Oct. 2021, doi: 10.1515/sjpain-2021-0060.

[72] M. J. L. Sullivan, “Toward a Biopsychomotor Conceptualization of Pain Implications for Research and Intervention,” 2008.

[73] B. Lindström and M. Eriksson, “Salutogenesis,” Journal of Epidemiology and Community Health, vol. 59, no. 6. pp. 440–442, Jun. 2005. doi: 10.1136/jech.2005.034777.

[74] DP. Ausubel, “The psychology of meaningful verbal learning.,” in New York: Grune & Stratton; 1963. 255 *p.,* 1963.

[75] T. G. K. Bryce and E. J. Blown, “Ausubel’s meaningful learning re-visited,” Current Psychology, 2023, doi: 10.1007/s12144-023-04440-4.

[76] D. P. Ausubel, Educational psychology: a cognitive view. 1968.

[77] A. Antonovxy, “THE STRUCTURE AND PROPERTIES OF THE SENSE OF COHERENCE SCALE,” 1993.

[78] M. Eriksson and B. Lindström, “Antonovsky’s sense of coherence scale and its relation with quality of life: A systematic review,” Journal of Epidemiology and Community Health, vol. 61, no. 11. pp. 938–944, Nov. 2007. doi: 10.1136/jech.2006.056028.

[79] P. M. Newton, A. Da Silva, and L. G. Peters, “A Pragmatic Master List of Action Verbs for Bloom’s Taxonomy,” Front Educ (Lausanne), vol. 5, Jul. 2020, doi: 10.3389/feduc.2020.00107.

[80] N. E. Adams, “Bloom’s taxonomy of cognitive learning objectives,” Journal of the Medical Library Association, vol. 103, no. 3, pp. 152–153, 2015, doi: 10.3163/1536-5050.103.3.010.

[81] Krathwohl DR, “From One Dimension to Two Dimensions,” 2002.

[82] K. D. Torralba and L. Doo, “Active Learning Strategies to Improve Progression from Knowledge to Action,” Rheumatic Disease Clinics of North America, vol. 46, no. 1. W.B. Saunders, pp. 1–19, Feb. 01, 2020. doi: 10.1016/j.rdc.2019.09.001.

[83] J. Phillips and F. Wiesbauer, “The flipped classroom in medical education: A new standard in teaching,” Trends in Anaesthesia and Critical Care, vol. 42, pp. 4–8, Feb. 2022, doi: 10.1016/j.tacc.2022.01.001.

[84] D. M. Higgins, A. M. Martin, D. G. Baker, J. J. Vasterling, and V. Risbrough, “The relationship between chronic pain and neurocognitive function a systematic review,” Clinical Journal of Pain, vol. 34, no. 3. Lippincott Williams and Wilkins, pp. 262– 275, Mar. 01, 2018. doi: 10.1097/AJP.0000000000000536.

[85] H. Ebbinghaus, “Memory: A Contribution to Experimental Psychology,” Ann Neurosci, vol. 20, no. 4, Oct. 2013, doi: 10.5214/ans.0972.7531.200408.

[86] J. Grisart, M. Van der Linden, and C. Bastin, “The contribution of recollection and familiarity to recognition memory performance in chronic pain patients,” Behaviour Research and Therapy, vol. 45, no. 5, pp. 1077–1084, May 2007, doi: 10.1016/j.brat.2006.05.002.

[87] P. A., & G. E. J. Woźniak, “Optimization of repetition spacing in the practice of learning.,” Acta Neurobiol Exp (Wars), pp. 54(1), 59–62., 1994.

[88] S. Fattinger et al., “Deep sleep maintains learning efficiency of the human brain,” Nat Commun, vol. 8, May 2017, doi: 10.1038/ncomms15405.

[89] W. R. Miller and S. Rollnick, Motivational interviewing: Preparing people to change addictive behavior., The Guilford Press. 1991.

[90] J. Nijs et al., “TITLE: Integrating Motivational Interviewing in Pain Neuroscience Education for People With Chronic Pain: A Practical Guide for Clinicians RUNNING HEAD: Motivational Interviewing Plus Explaining Pain TOC CATEGORY: Pain Management ARTICLE TYPE: Perspective”, doi: 10.1093/ptj/pzaa021/5716894.

[91] X. Bosch-Capblanch, K. Abba, M. Prictor, and P. Garner, “Contracts between patients and healthcare practitioners for improving patients’ adherence to treatment, prevention and health promotion activities,” Cochrane Database of Systematic Reviews, no. 2. 2007. doi: 10.1002/14651858.CD004808.pub3.

[92] F. Cuenca-Martínez, L. Suso-Martí, J. V. León-Hernández, and R. La Touche, “The role of movement representation techniques in the motor learning process: A neurophysiological hypothesis and a narrative review,” Brain Sciences, vol. 10, no. 1. MDPI AG, Jan. 01, 2020. doi: 10.3390/brainsci10010027.

